# Characteristics of 1,573 healthcare workers who underwent nasopharyngeal swab for SARS-CoV-2 in Milano, Lombardy, Italy

**DOI:** 10.1101/2020.05.07.20094276

**Authors:** Andrea Lombardi, Dario Consonni, Michele Carugno, Giorgio Bozzi, Davide Mangioni, Antonio Muscatello, Valeria Castelli, Emanuele Palomba, Anna Paola Cantù, Ferruccio Ceriotti, Basilio Tiso, Angela Cecilia Pesatori, Luciano Riboldi, Alessandra Bandera, Andrea Gori

## Abstract

**Background:** The management of healthcare workers (HCWs) exposed to confirmed cases of COVID-19 is still a matter of debate. It is unclear whether these subjects should be tested in the absence of symptoms and if those can guide diagnosis.

**Methods:** Occupational and clinical characteristics of all the consecutive HCWs who performed a nasopharyngeal swab for the detection of SARS-CoV-2 in a University Hospital from February 24, 2020, to March 31, 2020, were collected. Frequencies of positive tests were compared according to selected variables. Multivariable logistic regression analyses were then applied.

**Findings:** Positive tests were 138 among 1,573 HCWs (8.8%, 95% confidence interval [CI]: 7.4-10.3), with a marked difference between symptomatic (20.2%, 95% CI: 16.7-24.1) and asymptomatic (3.7%, 95% CI: 2.7-5.1) subjects (p<0.001). Physicians were the group with the highest frequency of positive tests (10.6%, 95% CI: 8.3-13.4) whereas clerical workers and technicians displayed the lowest frequency (2.9%, 95% CI: 0.8-7.3). The likelihood of being positive increased with the number of reported symptoms and the strongest predictors of a positive test were taste and smell alterations (odds ratio [OR] = 29.7) and fever (OR = 7.21). The median time from first positive test to a negative test was 23 days (95% CI: 19-24).

**Interpretation:** In this Italian group of HCWs exposed to confirmed cases of COVID-19 the presence of symptoms, especially taste and smell alterations and fever, was associated with SARS-CoV-2 infection. The median time to clear the virus from nasopharynx was 23 days.

**Funding:** none related to the content of this manuscript.

**Research in context:** *Evidence before this study:* We searched PubMed for articles published in English up to April 25, 2020, using the keywords “SARS-CoV-2”, “COVID-19”, “2019-nCoV”, AND “healthcare workers”,“HCW”, AND “testing”, “nasopharyngeal swab”. We found one article: *Roll-out of SARS-CoV-2 testing for healthcare workers at a large NHS Foundation Trust in the United Kingdom, March 2020* published in *Euro Surveillance*. Reviewing the pre-print website medRxiv with the same keywords we identified two additional studies: *SARS-CoV-2 infection in Health Care Workers in a large public hospital in Madrid, Spain, during March 2020*, and *SARS-CoV-2 infection in 86 healthcare workers in two Dutch hospitals in March*.

*Added value of this study:* We showed that, even if symptomatic healthcare workers had a much higher probability of positive test, almost one third of those infected were asymptomatic. Specific symptoms, namely taste and smell alterations and fever, were strongly associated with the infection. Finally, the median time to clear the virus from nasopharynx was 23 days.

*Implications of all the available evidence:* Screening strategies for healthcare workers exposed to COVID-19 patients should take in account the significant proportion of asymptomatic carriers and the predictive role of specific symptoms. Moreover, healthcare workers coming back to work after a positive test should be aware of the long-time of viral shedding from nasopharynx.

## Introduction

Severe acute respiratory syndrome coronavirus 2 (SARS-CoV-2) is a previously unknown virus which recently jumped from a not yet identified animal host to humans and it is responsible of coronavirus disease 2019 (COVID-19).^1^ This disease is characterized by a wide array of manifestations, ranging from an asymptomatic infection to a severe respiratory insufficiency requiring mechanical ventilatory support.^2^ The virus has now spread worldwide from China, causing the first pandemic of the XXI century, disrupting health-care services in the affected countries and exacting a terrific toll of human lives.^3–4^ A critical element of the virus is its basic reproduction number (R_0_) ranging from 2.76 to 3.28.^5,6^ This is the consequence of specific viral properties, the large number of asymptomatic, and thus undetected, carriers and the long duration of viral detectability, even after clinical cure.^7–10^ Currently, the only available method to ascertain the presence of SARS-CoV-2 infection is the detection of unique sequences of virus RNA by real-time reverse-transcription polymerase chain reaction (rRT-PCR) with confirmation by nucleic acid sequencing when necessary.^10^

Healthcare workers (HCWs) are a crucial actor of this pandemic with a Janus role. Indeed, they are acting in an emergency situation to mitigate the effects of the pandemic, but consequently they are continuously at risk of being infected. At the same time, they are in contact with the most fragile elements of our society, those who need health assistance. It is therefore mandatory to avoid that infected HCWs act as spreaders of the disease. Unfortunately, it is still unclear which microbiologic investigations and procedures should be adopted toward HCWs in COVID-19 settings, especially to those exposed to confirmed cases of COVID-19 and at risk for infection. To answer this question, we reviewed all the nasopharyngeal swab performed in HCWs exposed to confirmed cases of COVID-19 at the Foundation IRCCS Ca’Granda Ospedale Maggiore Policlinico located in Milan, the capital of Lombardy, by large the Italian region mostly affected by COVID-19.^11^ We assessed frequency of positive tests among symptomatic and asymptomatic subjects and evaluated the association between occupation, symptoms (type and number), and presence of the infection. Furthermore, we also calculated the median time between the day of diagnosis (first positive test) and the day in which the HCW became test-negative.

## Materials and methods

We collected occupational and clinical characteristics of all the consecutive HCWs who performed a nasopharyngeal swab for the detection of SARS-CoV-2 at the Foundation IRCCS Ca’Granda Ospedale Maggiore Policlinico in Milan, Italy in the period from February 24, 2020, (the day after the first COVID-19 case occurred in a physician of our hospital) to March 31, 2020. For these workers, we collected laboratory results as of April 9, 2020. We tested HCWs at risk for infection, which is defined as a contact with a patient or another HCW with (or later diagnosed with) SARS-CoV-2 infection. HCWs were subdivided into physicians (including residents), nurses and midwives, healthcare assistants, health technicians, and clerical workers and technicians. All the information was collected by the infectious disease notification form associated to each test. Subjects were defined as symptomatic if presented any of the following in the 14 days preceding the test: fever, cough, dyspnoea, asthenia, myalgia, coryza, sore throat, headache, ageusia or dysgeusia, anosmia or parosmia, ocular symptoms, diarrhoea, nausea, and vomit. The study was approved by the Ethical Committee of our institution and was conducted in accordance with the Helsinki Declaration.

### SARS-CoV-2 detection

For viral detection two different methods were used. The first one employed Seegene Inc reagents (Seoul, Korea). RNA extraction was performed with STARMag Universal Cartridge kit on Nimbus instrument (Hamilton, Agrate Brianza, Italy) and amplification with Allplex® 2019-nCoV assay. The second one employed a GeneFinder® COVID-19 Plus RealAmp Kit (OSANG Healthcare, Anyangcheondong-ro, Dongan-gu, Anyang-si, Gyeonggi-do, Korea) on ELITech InGenius® instrument (Torino, Italy). Both assays identify the virus by multiplex rRT-PCR targeting three viral genes (E, RdRP and N).

### Statistical analysis

We compared frequencies of positive tests according to selected variables using chi-squared test, adjusted odds ratios (OR), and 95% confidence intervals (CI) calculated with a multivariable logistic regression model including as covariates gender, age class, occupation, and having reported any symptom. We evaluated the discriminating ability of the number of reported symptoms in a univariate logistic model and assessed the performance of each of 11 groups of symptoms by fitting a multivariable logistic model containing all groups of symptoms. Area under the ROC curve (AUC) was calculated after these models. We calculated the time since first positive test until subjects became negative by using the Kaplan-Meier function. Log-rank test was used to evaluate the association of gender, age class, or symptoms with median time to test negativity. Statistical analysis was performed with Stata 16 (StataCorp. 2019)

## Results

In the period from February 24, 2020, to March 31, 2020, 1,573 HCWs, 1,010 women (64.2%) and 563 men (35.8%) performed at least a first nasopharyngeal swab for the detection of SARS-CoV-2. Mean age was 44.5 years and the majority (about 70%) were physicians (including residents) or nurses/midwives (table 1). One third of women and one fourth of men reported having had at least one symptom at the time of testing. The majority (73.9%) performed only one test, while 411 individuals (26.1%) had from two to six tests.

**Table 1.** Characteristics of 1,573 healthcare workers tested for SARS-CoV-2 in, Milan, Italy, in the period from February 24, 2020, to March 31, 2020.

|  | **Women** |  | **Men** |  | **Total** |  |
| --- | --- | --- | --- | --- | --- | --- |
| **Variable** | N | % | N | % | N | % |
| All subjects | 1010 | 100 | 563 | 100 | 1573 | 100 |
| Age (years), mean (min-max) | 44.2 | (21-67) | 45.1 | (22-76) | 44.5 | (21-76) |
| Age (years) |  |  |  |  |  |  |
| <30 | 152 | 15.1 | 96 | 17.0 | 248 | 15.8 |
| 30-39 | 255 | 25.2 | 132 | 23.5 | 387 | 24.6 |
| 40-49 | 217 | 21.5 | 109 | 19.4 | 326 | 20.7 |
| 50-59 | 314 | 31.1 | 130 | 23.1 | 444 | 28.2 |
| 60+ | 72 | 7.1 | 96 | 17.0 | 168 | 10.7 |
| Occupation |  |  |  |  |  |  |
| Physicians, including residents | 295 | 29.2 | 287 | 51.0 | 582 | 37 |
| Nurses, midwives | 388 | 38.4 | 134 | 23.8 | 522 | 33.2 |
| Healthcare assistants | 122 | 12.1 | 40 | 7.1 | 162 | 10.3 |
| Health technicians* | 133 | 13.2 | 37 | 6.6 | 170 | 10.8 |
| Clerical workers, technicians | 72 | 7.1 | 65 | 11.5 | 137 | 8.7 |
| At least one symptom | 343 | 34.0 | 137 | 24.3 | 480 | 30.5 |
| No. tests performed |  |  |  |  |  |  |
| 1 | 759 | 75.1 | 403 | 71.6 | 1162 | 73.9 |
| 2 | 161 | 15.9 | 99 | 17.6 | 260 | 16.5 |
| 3 | 59 | 5.8 | 40 | 7.1 | 99 | 6.3 |
| 4 | 19 | 1.9 | 12 | 2.1 | 31 | 2.0 |
| 5 | 9 | 0.9 | 3 | 0.5 | 12 | 0.8 |
| 6 | 3 | 0.3 | 6 | 1.1 | 9 | 0.6 |

\*Includes biologists, radiology and laboratory technicians, psychologists, other health technicians

The overall frequency of subjects with at least one positive test was 8.8% (95% CI: 7.4-10.3%) (table 2). The frequency of positive tests ranged from 8.0% (healthcare assistants) to 10.6% (physicians), much higher than among clerical workers (2.9%). Among subjects with symptoms the frequency of positive tests was 20.2%, while among asymptomatic HCWs the frequency was much lower (3.7%). However, among the 138 HCWs with positive test, 41/138 (29.7%) were asymptomatic. The predictive role of occupation and presence of symptoms was confirmed in the multivariable logistic model.

**Table 2.**
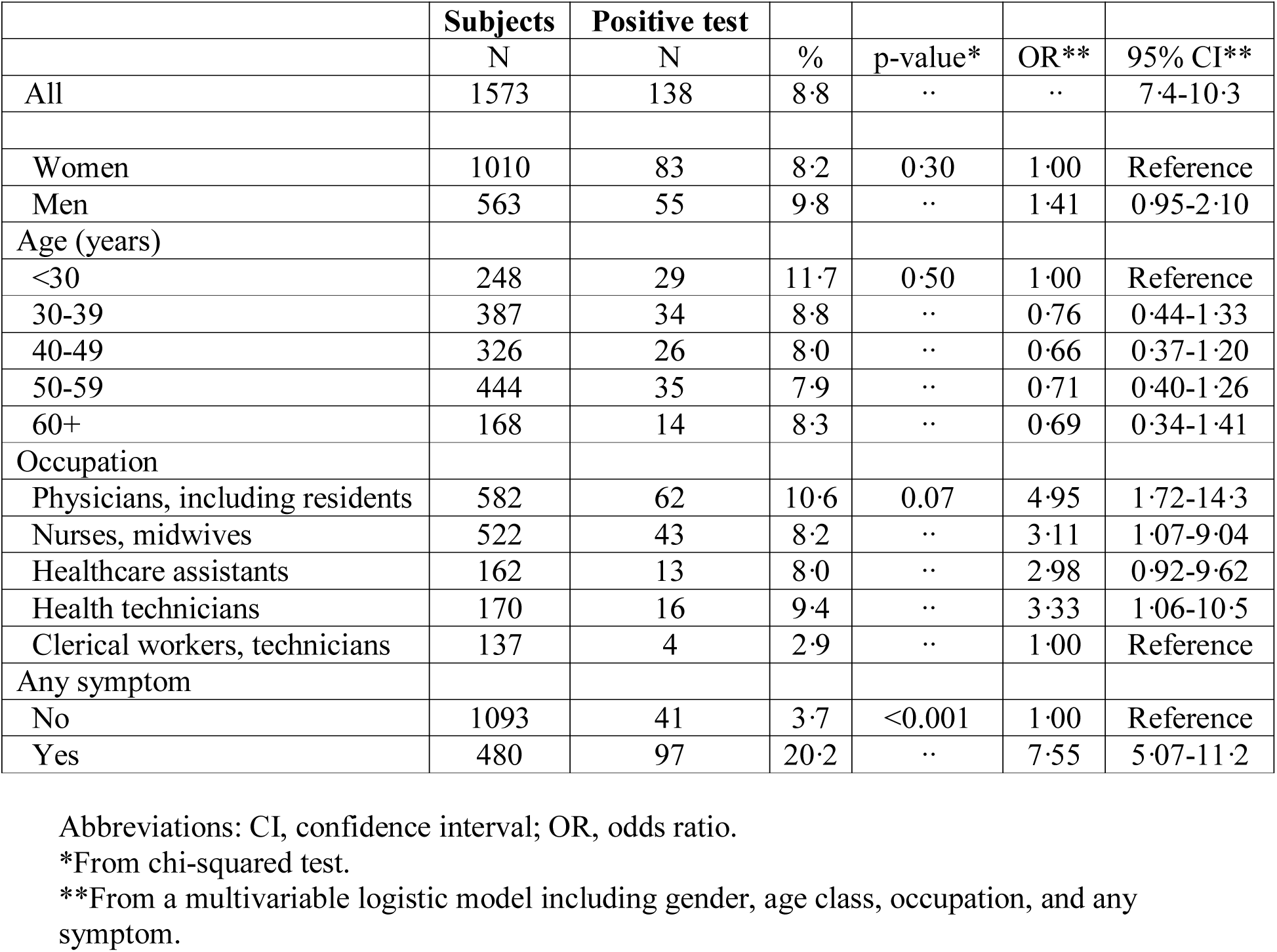
Association between selected variables and frequency of at least one positive test among 1,573 healthcare workers tested for SARS-CoV-2 in Milan, Italy, in the period from February 24, 2020, to March 31, 2020.

|  | **Subjects** | **Positive test** |  |  |  |  |
| --- | --- | --- | --- | --- | --- | --- |
|  | N | N | % | p-value* | OR** | 95% CI** |
| All | 1573 | 138 | 8.8 | .. | .. | 7.4-10.3 |
| Women | 1010 | 83 | 8.2 | 0.30 | 1.00 | Reference |
| Men | 563 | 55 | 9.8 | .. | 1.41 | 0.95-2.10 |
| Age (years) |  |  |  |  |  |  |
| <30 | 248 | 29 | 11.7 | 0.50 | 1.00 | Reference |
| 30-39 | 387 | 34 | 8.8 | .. | 0.76 | 0.44-1.33 |
| 40-49 | 326 | 26 | 8.0 | .. | 0.66 | 0.37-1.20 |
| 50-59 | 444 | 35 | 7.9 | .. | 0.71 | 0.40-1.26 |
| 60+ | 168 | 14 | 8.3 | .. | 0.69 | 0.34-1.41 |
| Occupation |  |  |  |  |  |  |
| Physicians, including residents | 582 | 62 | 10.6 | 0.07 | 4.95 | 1.72-14.3 |
| Nurses, midwives | 522 | 43 | 8.2 | .. | 3.11 | 1.07-9.04 |
| Healthcare assistants | 162 | 13 | 8.0 | .. | 2.98 | 0.92-9.62 |
| Health technicians | 170 | 16 | 9.4 | .. | 3.33 | 1.06-10.5 |
| Clerical workers, technicians | 137 | 4 | 2.9 | .. | 1.00 | Reference |
| Any symptom |  |  |  |  |  |  |
| No | 1093 | 41 | 3.7 | <0.001 | 1.00 | Reference |
| Yes | 480 | 97 | 20.2 | .. | 7.55 | 5.07-11.2 |

Abbreviations: CI, confidence interval; OR, odds ratio. \*From chi-squared test. \*\*From a multivariable logistic model including gender, age class, occupation, and any symptom.

The likelihood of being positive increased with the number of reported symptoms (table 3). All symptoms excluding sore throat were positively associated with test positivity, especially fever and taste and smell alterations (table 4). In a multivariable model, the strongest predictors of a positive test were taste and smell alterations (OR = 29.7) and fever (OR = 7.21), followed by myalgias, asthenia, ocular symptoms, and dyspnoea (ORs ranging from 1.98 and 2.77). The AUC from the model including these six group of symptoms was 0.74 (95% CI: 0.70-0.79), similar to an AUC of 0.77 (95% CI: 0.72-0.81) when including all symptoms. Sore throat was negatively associated with positivity (OR = 0.35).

**Table 3.** Association between number of symptoms and frequency of at least one positive test among 1,573 healthcare workers tested for SARS-CoV-2 in Milan, Italy, in the period from February 24, 2020, to March 31, 2020.

|  | **Subjects** | **Positive test** |  |  |  |  |
| --- | --- | --- | --- | --- | --- | --- |
|  | N | N | % | p-value* | OR** | 95% CI** |
| **Number of symptoms** |  |  |  |  |  |  |
| No symptoms | 1093 | 41 | 3.7 | <0.001 | 1.00 | Reference |
| 1 | 191 | 26 | 13.6 | .. | 4.04 | 2.41-6.79 |
| 2 | 145 | 29 | 20.0 | .. | 6.41 | 3.84-10.7 |
| 3 | 98 | 27 | 27.8 | .. | 9.76 | 5.67-16.8 |
| 4 | 35 | 10 | 28.6 | .. | 10.3 | 4.63-22.8 |
| 5 | 7 | 3 | 42.8 | .. | 19.2 | 4.17-88.8 |
| 6 | 4 | 2 | 50.0 | .. | 25.7 | 3.53-186 |

Abbreviations: CI, confidence interval; OR, odds ratio.
\*From chi-squared test.
\*\*From a univariate logistic model.

**Table 4.**
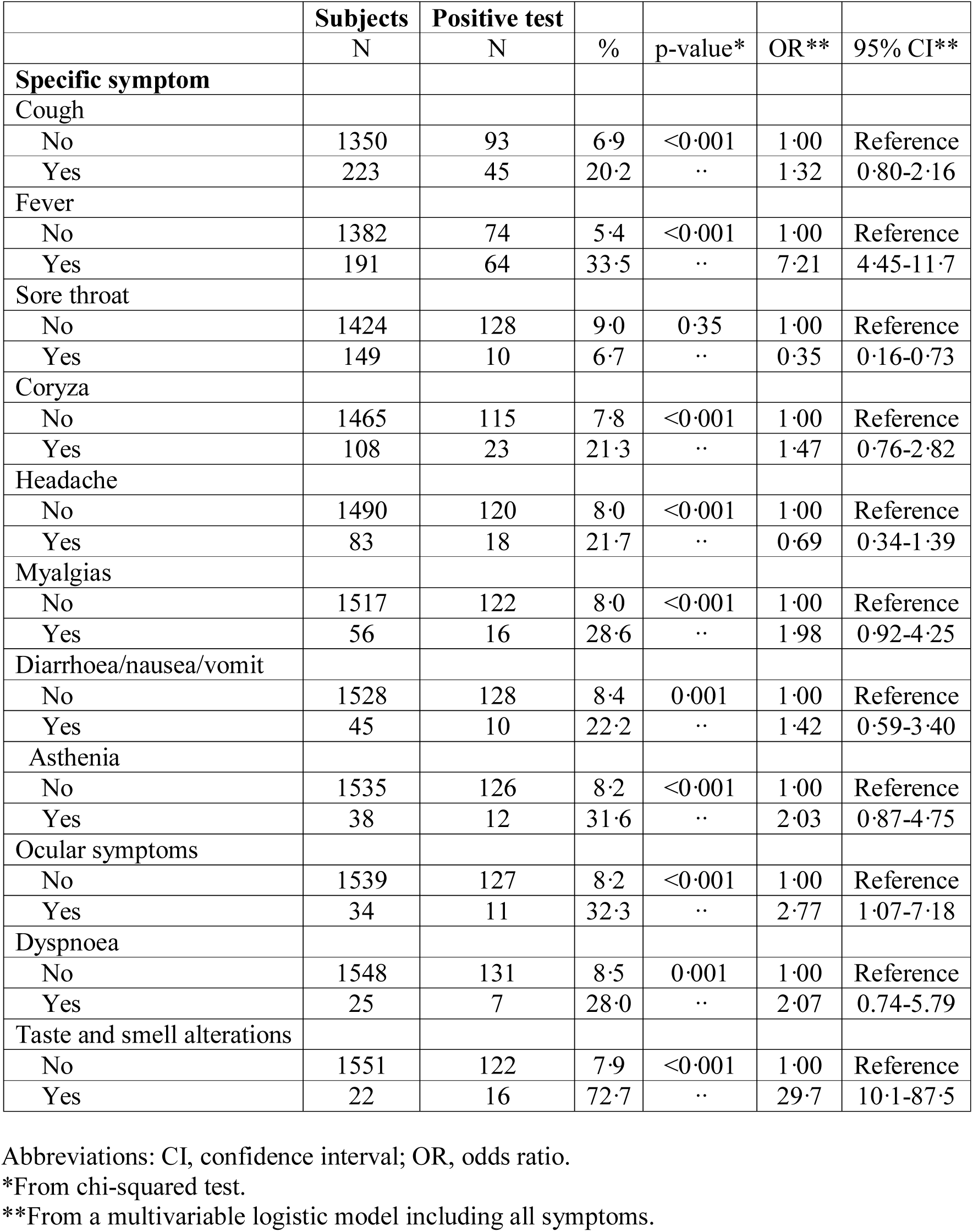
Association between selected symptoms and frequency of at least one positive tests among 1,573 healthcare workers tested for SARS-CoV-2 in Milan, Italy, in the period from February 24, 2020, to March 31, 2020.

|  | Subjects | Positive test |  |  |  |  |
| --- | --- | --- | --- | --- | --- | --- |
|  | N | N | % | p-value* | OR** | 95% CI** |
| **Specific symptom** |  |  |  |  |  |  |
| Cough |  |  |  |  |  |  |
| No | 1350 | 93 | 6.9 | <0.001 | 1.00 | Reference |
| Yes | 223 | 45 | 20.2 | .. | 1.32 | 0.80-2.16 |
| Fever |  |  |  |  |  |  |
| No | 1382 | 74 | 5.4 | <0.001 | 1.00 | Reference |
| Yes | 191 | 64 | 33.5 | .. | 7.21 | 4.45-11.7 |
| Sore throat |  |  |  |  |  |  |
| No | 1424 | 128 | 9.0 | 0.35 | 1.00 | Reference |
| Yes | 149 | 10 | 6.7 | .. | 0.35 | 0.16-0.73 |
| Coryza |  |  |  |  |  |  |
| No | 1465 | 115 | 7.8 | <0.001 | 1.00 | Reference |
| Yes | 108 | 23 | 21.3 | .. | 1.47 | 0.76-2.82 |
| Headache |  |  |  |  |  |  |
| No | 1490 | 120 | 8.0 | <0.001 | 1.00 | Reference |
| Yes | 83 | 18 | 21.7 | .. | 0.69 | 0.34-1.39 |
| Myalgias |  |  |  |  |  |  |
| No | 1517 | 122 | 8.0 | <0.001 | 1.00 | Reference |
| Yes | 56 | 16 | 28.6 | .. | 1.98 | 0.92-4.25 |
| Diarrhoea/nausea/vomit |  |  |  |  |  |  |
| No | 1528 | 128 | 8.4 | 0.001 | 1.00 | Reference |
| Yes | 45 | 10 | 22.2 | .. | 1.42 | 0.59-3.40 |
| Asthenia |  |  |  |  |  |  |
| No | 1535 | 126 | 8.2 | <0.001 | 1.00 | Reference |
| Yes | 38 | 12 | 31.6 | .. | 2.03 | 0.87-4.75 |
| Ocular symptoms |  |  |  |  |  |  |
| No | 1539 | 127 | 8.2 | <0.001 | 1.00 | Reference |
| Yes | 34 | 11 | 32.3 | .. | 2.77 | 1.07-7.18 |
| Dyspnoea |  |  |  |  |  |  |
| No | 1548 | 131 | 8.5 | 0.001 | 1.00 | Reference |
| Yes | 25 | 7 | 28.0 | .. | 2.07 | 0.74-5.79 |
| Taste and smell alterations |  |  |  |  |  |  |
| No | 1551 | 122 | 7.9 | <0.001 | 1.00 | Reference |
| Yes | 22 | 16 | 72.7 | .. | 29.7 | 10.1-87.5 |

Abbreviations: CI, confidence interval; OR, odds ratio.
\*From chi-squared test.
\*\*From a multivariable logistic model including all symptoms.

Among the 138 positive HCWs, 99 (71.7%) were already positive at first testing, while 39 (28.3%) were found positive in a subsequent test. At the time of last test performed, 69/138 (50.0%) were still positive. The median time from first positive test to a negative test was 23 days (95% CI: 19-24) (figure 1). However, 9/69 subjects (13.0%) were still positive 24 to 31 days since first positive test. Median time was identical in subjects with symptoms (23 days, 95% CI: 18-26) and in those without symptoms (23 days, 95% CI: 19.29). Median time was also not associated with gender (p=0.84) nor age (p=0.83). As of March 31, 2020, five workers, four men (three physicians and a nurse) and a woman (clerical worker) were hospitalized.

**Figure 1.**
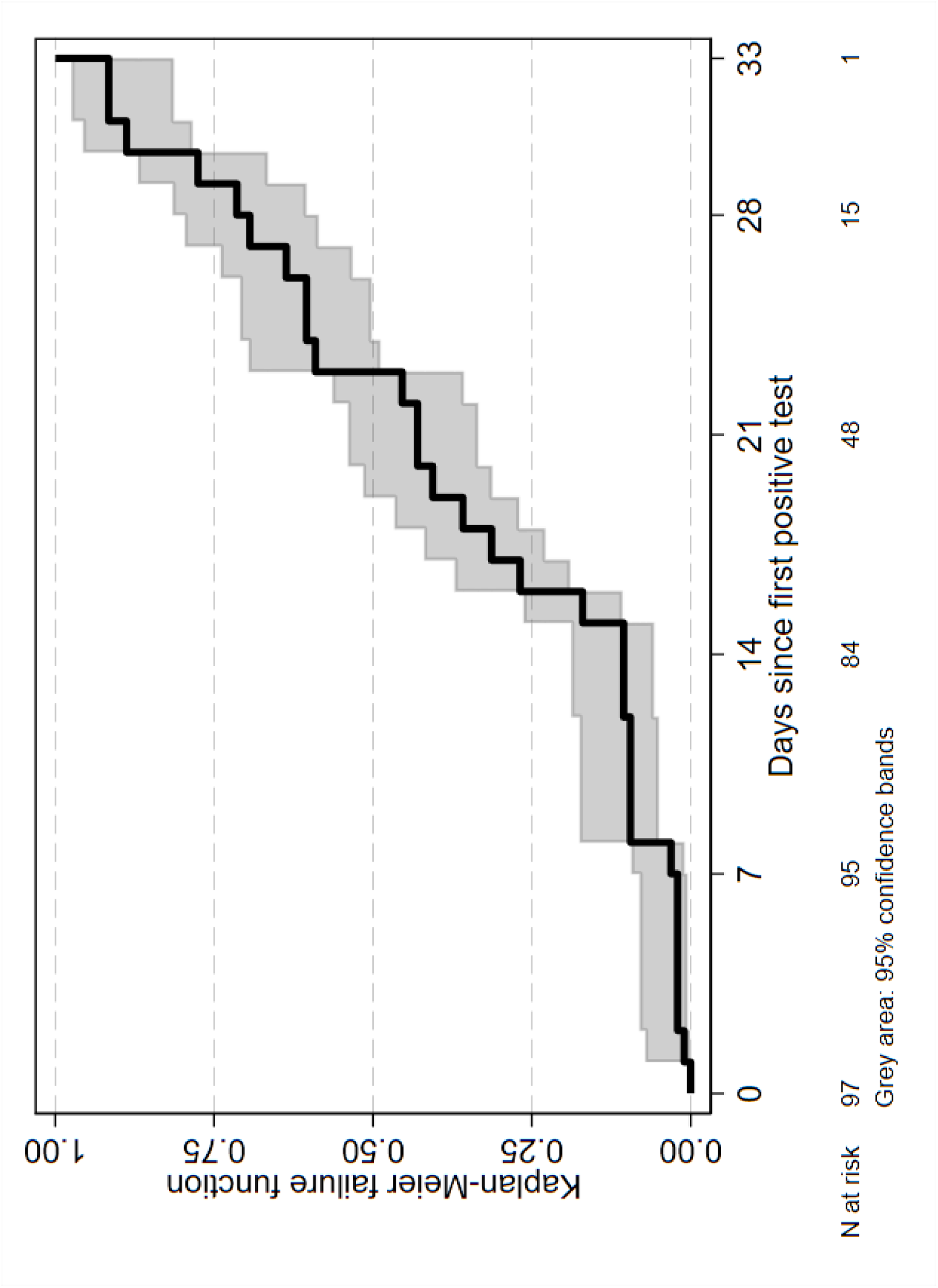
Kaplan-Meier failure function showing times at which subjects became test-negatives.

A minority of the HCWs (81/1,537, 5.3%) reported to have had a contact with an infected person outside the hospital (relatives, colleague, or friends). Of these, 12 (8.7%) were found to be positive.

## Discussion

In this Italian group of HCWs exposed to confirmed cases of COVID-19, the presence of symptoms, and particularly taste and smell alterations and fever, was associated with positivity of nasopharyngeal swab for SARS-CoV-2. Despite the low relative frequency of positive tests among asymptomatic subjects, their number was high in absolute terms (just about one third of all infected subjects). Interestingly, the AUC of a model considering six groups of symptoms (fever, myalgia, asthenia, ocular symptoms, dyspnoea, and taste and smell alterations) was 0.74. Based on these results, it seems reasonable to tailor the screening approach of HCWs at risk based on the resources available. In low-resource settings we suggest focusing to test those with symptoms to maximize efficacy, especially considering the continuous exposure of HCWs to at risk situations, thus requiring repeated testing sessions. Nevertheless, it should be underlined that in our study 41/138 subjects (29.7%) were infected but displayed no symptoms, meaning that one third of those infected can be lost with a symptoms-based screening strategy. Therefore, in middle- and high-resource settings a mass screening for all HCWs exposed to confirmed COVID-19 cases appears the best approach to limit the spread of the virus. More detailed cost-effectiveness study, encompassing the epidemiological context, should be performed to define the optimal method.

The frequency of positive subjects among symptomatic workers in our study population (20.2%) is similar to the one (18%) reported by Keeley and colleagues^12^ in their cohort composed of 1,533 symptomatic HCWs presenting with fever plus one among cough, sore throat, runny nose, myalgia, headache and persistent cough. However, it should be remarked that focussing only on symptomatic workers results in missing a significant number of infected subjects. Indeed, we had 67/138 (48.5%) positive HCWs presenting without or with only one symptom. When we consider the overall frequency, our proportion of positive subjects (8.8%) is comparable to the 6% described by Kluytmans-van den Bergh et al. in a small Dutch cohort of HCWs, whereas it is significantly lower than the 38% reported by Folgueira and colleagues in their Spanish cohort.^13,14^

When stratified according to occupation, test-positive frequencies were clearly higher among subsets with direct contact with patients (physicians including residents, nurses and midwives, healthcare assistants and health technicians) than those without (clerical works and technicians). Consequently, careful screening of these groups of workers should be mandatory. No differences in terms of infection prevalence were seen between different age groups nor between men and women, suggesting that risk factors for acquiring COVID-19 among HCWs are unrelated to age and sex.

Another relevant point is the significant number of subjects who were negative at the first test but resulted positive when tested a second time. This might represent a serious concern, as a discrete fraction of those can further spread the virus unnoticed, thus hampering the efficacy of the screening strategy. It should be noted, however, that the second test was performed on a small number of operators and not on a routine basis, making these considerations subject to several potential biases. In addition, in a relevant proportion of our population we could not retrieve information about the most likely date of exposure to a documented COVID-19 case. Thus, we cannot exclude a recent contact in which case the first test may have been performed too early (i.e. still in the incubation period which has been estimated to be five days), before a sufficient amount of viral particles is detectable in the nasopharynx.^15^

Moreover, it has to be considered that HCWs employed in COVID-19 units/hospitals are at risk of SARS-CoV-2 exposure on a daily basis and therefore repeated exposures, even unnoticed, can occur also after the first one who motivated the test. Moreover, technical limitation can be responsible of falsely negative test, considering that the sensitivity of nasopharyngeal swab for SARS-CoV-2 detection has been estimated to be around 71%.^16^

Finally, even if it was not the main goal of our study (because a longer follow-up time would be required), we observed a median time from first positive test to a negative test of 23 days. This is in accordance with several already published reports and have a significant impact on the efficiency of health systems.^17–19^ Indeed, it means that an infected HCW will be unavailable to perform its duty for at least three weeks since diagnosis (or even four weeks if we consider the upper 95% confidence limit). Our results were based on a three genes qualitative RT-PCR. To understand the real significance of this viral detection new studies assessing the infectivity of viral particles and the possible impact of quantitative techniques are needed.

In conclusion, our results show that symptomatic HCWs exposed to confirmed cases of COVID-19 are almost eight times more likely to be infected than asymptomatic HCWs. Nevertheless, also a non-negligible amount of asymptomatic HCWs is infected and accounts for almost one third of positive tests. Therefore, screening strategies may be tailored according to the available resources. Taste and smell alterations and fever should be considered the most relevant alarm bells suggesting the opportunity of performing a test. Finally, the median time to become non-infective exceeded three weeks. Consequently, the suggested quarantine period of 14 days after exposure to a confirmed case should be revised. The correct length of this period as well as the best moment to perform a nasopharyngeal swab (measured in days after exposure) have to to be determined.

## Data Availability

Data will be available on demand

## Authors contributions

AL, DC and AG conceived the study. APC, BT, VC, EP and LR collected the data. DC, MC, and ACP performed statistical analyses. AL and DC wrote the first draft. All co-authors revised the manuscript.

## Acknowledgments

We thank the personnel of the Virology Unit and SPIO (Servizio Prevenzione e Igiene Ospedaliera), Francesco De Palo, Lidia Guerrieri, Lorenzo Bordini, Carlo Nava, Antonio Lela, Enrico Radice, and Carolina Mensi for their help in data collection and editing. The authors are also grateful to Simone Villa for the critical review of the manuscript.

